# Patient and caregiver lived experiences and mental health service engagement during first-episode psychosis in Uganda: a longitudinal mixed-methods study protocol

**DOI:** 10.64898/2026.05.21.26353795

**Authors:** Prisca Oroma, Andrew Ssentoogo Ssemata, Wilber Ssembajjwe, Rita Auma, Sophia Balinga, Blessed Tabitha Aujo, Andrea Kaggwa Kaddu, Mary Ampiire, Wilson Winstons Muhwezi, Emmanuel Kiiza Mwesiga, Etheldreda Nakimuli

## Abstract

**Introduction:** Engagement with mental health services (MHCS) during the first episode of psychosis (FEP) is critical for symptom control, quality of life, and relapse prevention. However, disengagement rates remain high in Uganda with severe consequences for patients and caregivers. This study protocol describes a mixed-methods investigation which aims to examine the relationship between patients and caregivers lived experiences and mental health service engagement during first-episode psychosis.

**Methods and Analysis:** The mixed-methods study will recruit 82 patients with first-episode psychosis and their primary caregivers from Butabika National Referral Mental Hospital in Kampala, Uganda. Inclusion criteria are ages 18-60, less than 12 weeks on antipsychotic medications, living in the greater Kampala Metropolitan Area, with a consenting caregiver. Caregivers must be an adult (> 18years) providing full-time care for at least 6 months prior. Patients with substance use disorders will be excluded. Qualitative data on the lived experiences of patients and caregivers will be collected using the draw-write-and-tell method, while quantitative data on service engagement and associated factors will be collected using semi-structured questionnaires. The data will be analysed using Stata version 18, and participants will be reimbursed for their time.

**Ethics and Dissemination:** Ethical clearance has been obtained from the School of Medicine Research and Ethics Committee (SOMREC) Ref: Mak-SOMREC-2024-1002 and institutional approval from Butabiika National Referral Mental Hospital. All participants will provide informed consent prior to participation. Data will be de-identified and securely stored, with results disseminated through peer-reviewed academic publications, conferences and community stakeholder workshops.

**Strength and limitations of the study:**

▯ This longitudinal mixed-methods study integrates patient and caregiver perspectives, enabling a holistic understanding of mental health service engagement during the first episode of psychosis. Particularly the use of an innovative draw-write-and-tell method for qualitative research in this population is novel.
▯ Inclusion of lived-experience narratives alongside service engagement trajectories strengthens the relevance of findings for mental health service design and early intervention strategies in low-resource settings.
▯ Conducting the study at a national referral mental health hospital allows in-depth examination of care pathways within a specialist service context; however, findings may not be fully generalisable to community-based or non-specialist settings.
▯ Exclusion of individuals with comorbid substance use disorders may limit the applicability of findings, given the high prevalence of substance use among people experiencing first-episode psychosis.

## Background

Early engagement with mental health care services during the first episode of psychosis (FEP) is consistently associated with better symptomatic and functional outcomes, including improved quality of life, reduced disability, and lower mortality (1, 2). However, continued engagement with mental health care services after first contact is often discontinuous, with cycles of disengagement and re-entry that effectively extend the duration of untreated or inadequately treated illness (3–5). In Uganda, these challenges are particularly pronounced. Early pathways to care are frequently shaped by caregivers who initially pursue alternative and complementary therapies before reaching formal services (2, 6, 7). Patients commonly present late and severely unwell, but even after inpatient stabilisation, disengagement with mental health services is as high as 70%, with high rates of failure to return for follow-up and measurable excess mortality among those who disengage (2, 6). The benefits of early psychosis care are only realised when patients and families can enter and remain in care early enough and long enough for those treatments to change long-term trajectories.

What remains unclear is why engagement is so unstable in the Ugandan FEP context, and which levers are most actionable for improving continuity of care. Existing studies have described disengagement rates and associated outcomes (2, 6), but their predictors have limited generalisability. The mechanisms by which engagement is produced or lost remain unclear. In particular, the role of patient and caregiver lived experience has not been sufficiently examined. Recent syntheses of lived experiences during FEP have identified common thematic domains, but they have largely excluded low-income countries and have not examined how these experiences map onto concrete engagement outcomes (discharge follow-up, retention, and re-engagement) (6, 8–10). It is unclear how patients and their caregivers interpret symptoms, experience stigma, build trust with help systems or how early encounters with both alternative and formal providers shape subsequent choices. As a result, the field lacks a context-sensitive account of how the lived experience of patients and caregivers’ shapes engagement during FEP in Uganda.

### Aims

This study aims to address this gap by examining patient and caregiver lived experiences during FEP and their relationship to engagement with mental health services in Uganda. Specifically, we aim to 1) explore patient and caregiver lived experiences during the first episode of psychosis in Uganda, 2) examine mental healthcare services engagement during the first episode of psychosis in Uganda and 3) examine the link between patient and caregiver lived experiences with mental health care services engagement during the first episode of psychosis in Uganda. Understanding these experiences will inform the design of culturally appropriate interventions to improve early engagement, reduce the duration of untreated psychosis, and enhance clinical and functional outcomes for both patients and caregivers.

## METHODS AND ANALYSIS

### Study Design

This study will employ a prospective longitudinal mixed-methods design to examine how patient and caregiver lived experiences influence mental health service engagement during the first episode of psychosis (FEP). A mixed-methods approach is appropriate as service engagement is shaped by both measurable clinical and service-related factors and contextual, experiential, and relational processes that are best captured qualitatively (11). Quantitative and qualitative data will be collected from patients and their caregivers at three time points: at enrolment in care (baseline) and at 3- and 6-month follow-up clinic visits. The quantitative and qualitative components will be integrated at the interpretation stage to generate complementary insights into patterns and processes of service engagement. The longitudinal design will allow examination of changes in experiences and patterns of service engagement over time and how these evolve during early contact with mental health services.

### Study Setting and site

The study will be conducted at Butabika National Referral Mental Hospital located approximately 13 km east of Kampala, Uganda. Butabika is Uganda’s main national referral psychiatric hospital and plays a central role in the delivery of specialist mental health services, training, and policy direction. The hospital receives patients from across the country and serves as the primary referral centre for individuals experiencing their first episode of psychosis. The research team has prior experience recruiting and following patients with FEP at this site, supporting the feasibility of longitudinal follow-up (2, 5–7, 12–17). As a national referral facility, Butabika provides an appropriate setting for exploring patients’ and caregivers’ lived experiences of mental health care engagement within Uganda’s public mental health system.

### Participants

Participants will be recruited from patients attending the Psychosis Clinic at Butabika National Referral Mental Hospital. Enrolment is open to all patients reporting to the clinic and diagnosed with first-episode psychosis. Eligible patients will be identified during routine clinical care and approached by trained research staff for study participation. Caregivers of participating patients will also be invited to take part in the study, contingent on patient consent. Recruitment commenced in 2025 and is expected to continue until June 2026, with longitudinal follow-up conducted at 3- and 6-months post-enrolment.

### Participant Eligibility Criteria

Patients will be eligible for inclusion if they meet all of the following criteria: a) are 18 – 50 years of age; b) have a diagnosis of a psychotic disorder confirmed using the Mini International Neuropsychiatric Interview (MINI), a validated structured diagnostic interview (18); c) are experiencing a first episode of psychosis, defined as first admission for a psychotic disorder; d) have received antipsychotic medication for less than 12 weeks at the time of enrolment (19, 20); e) are admitted to the convalescent wards of Butabika Hospital; f) reside within the Greater Kampala Metropolitan Area, to facilitate follow-up; and g) have a caregiver willing to participate in the study. Patients with a current substance use disorder as determined by the MINI will be excluded. This exclusion is intended to reduce clinical heterogeneity and allow focused examination of mental health service engagement trajectories among individuals with first episode psychosis (21).

Caregivers will be eligible for inclusion if they: are the primary caregiver responsible for the participating patient; a) are aged 18 years or older; b) have lived with the patient full-time for at least six months prior to hospital admission; c) provide written informed consent to participate.

### Study procedure for the Quantitative component

#### Sampling and recruitment

Butabika National Referral Mental Hospital is the national psychiatric referral hospital (15). Given this context, the study will recruit a consecutive sample of eligible patients presenting to the psychosis clinic and admitted to the convalescent wards during the recruitment period. Members of the research team with clinical experience in managing patients with first-episode psychosis will be trained in study procedures. In collaboration with the hospital records team, the research team will screen admission registers to identify patients who meet the eligibility criteria. Eligible patients will be approached during their admission to receive information about the study, undergo eligibility confirmation, and provide written informed consent. Following patient consent, caregivers will be contacted to assess their willingness to participate. Caregivers who consent will be invited to complete baseline assessments, typically on or near the patient’s day of discharge.

#### Baseline assessments

At enrolment (baseline), quantitative data will be collected from patients and caregivers to document socio-demographic, clinical, and psychosocial characteristics using standardized measures (Table 1). These baseline variables will serve as potential predictors of subsequent mental health care service (MHCS) engagement.

**Table 1:**
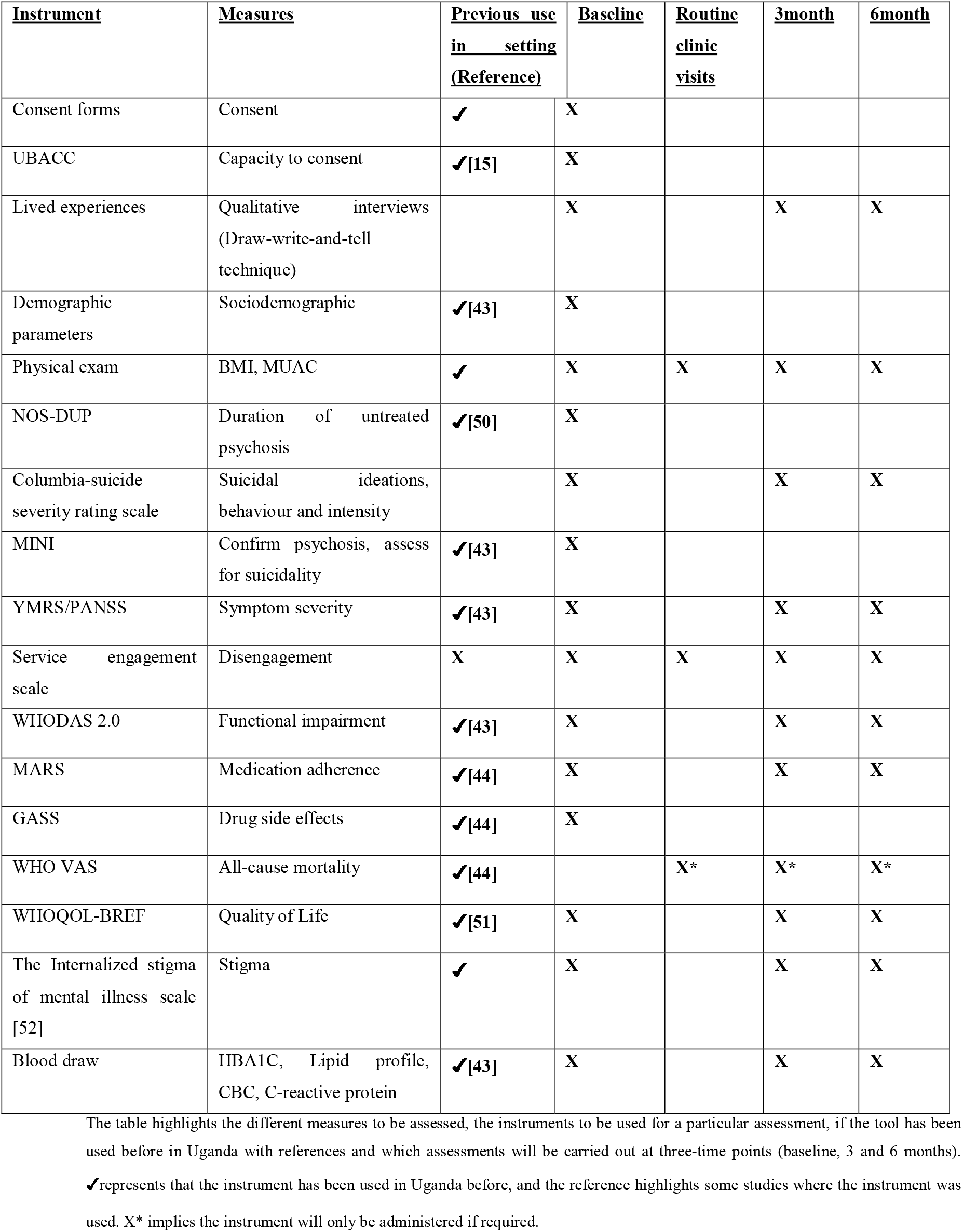
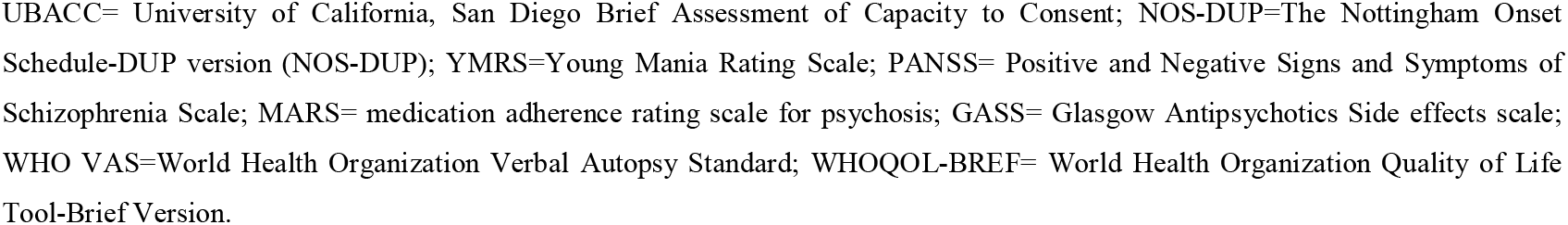
Study measures.

| <u>**Instrument**</u> | <u>**Measures**</u> | <u>**Previous use**</u><br><u>**in setting**</u><br><u>**(Reference)**</u> | <u>**Baseline**</u> | <u>**Routine**</u><br><u>**clinic**</u><br><u>**visits**</u> | <u>**3month**</u> | <u>**6month**</u> |
| --- | --- | --- | --- | --- | --- | --- |
| Consent forms | Consent | ✓ | X |  |  |  |
| UBACC | Capacity to consent | ✓[15] | X |  |  |  |
| Lived experiences | Qualitative interviews<br>(Draw-write-and-tell<br>technique) |  | X |  | X | X |
| Demographic<br>parameters | Sociodemographic | ✓[43] | X |  |  |  |
| Physical exam | BMI, MUAC | ✓ | X | X | X | X |
| NOS-DUP | Duration of untreated<br>psychosis | ✓[50] | X |  |  |  |
| Columbia-suicide<br>severity rating scale | Suicidal ideations,<br>behaviour and intensity |  | X |  | X | X |
| MINI | Confirm psychosis, assess<br>for suicidality | ✓[43] | X |  |  |  |
| YMRS/PANSS | Symptom severity | ✓[43] | X |  | X | X |
| Service engagement<br>scale | Disengagement | X | X | X | X | X |
| WHODAS 2.0 | Functional impairment | ✓[43] | X |  | X | X |
| MARS | Medication adherence | ✓[44] | X |  | X | X |
| GASS | Drug side effects | ✓[44] | X |  |  |  |
| WHO VAS | All-cause mortality | ✓[44] |  | X* | X* | X* |
| WHOQOL-BREF | Quality of Life | ✓[51] | X |  | X | X |
| The Internalized stigma<br>of mental illness scale<br>[52] | Stigma | ✓ | X |  | X | X |
| Blood draw | HBA1C, Lipid profile,<br>CBC, C-reactive protein | ✓[43] | X |  | X | X |

The table highlights the different measures to be assessed, the instruments to be used for a particular assessment, if the tool has been used before in Uganda with references and which assessments will be carried out at three-time points (baseline, 3 and 6 months).
✓ represents that the instrument has been used in Uganda before, and the reference highlights some studies where the instrument was used. X\* implies the instrument will only be administered if required.
UBACC= University of California, San Diego Brief Assessment of Capacity to Consent; NOS-DUP=The Nottingham Onset Schedule-DUP version (NOS-DUP); YMRS=Young Mania Rating Scale; PANSS= Positive and Negative Signs and Symptoms of Schizophrenia Scale; MARS= medication adherence rating scale for psychosis; GASS= Glasgow Antipsychotics Side effects scale; WHO VAS=World Health Organization Verbal Autopsy Standard; WHOQOL-BREF= World Health Organization Quality of Life Tool-Brief Version.

#### Follow-up and outcome assessment

After discharge, study staff will liaise with the hospital records department to track attendance at scheduled outpatient review visits. MHCS engagement will be assessed using the Service Engagement Scale (SES), a clinician-rated measure of patient and caregiver engagement with mental health services (22). Participants who attend their scheduled review visits will be assessed using the SES during routine clinic encounters.

Participants who fail to attend a scheduled review within one week of the appointment date will be assigned an SES score of zero for that time point, indicating non-engagement. Follow-up assessments will be conducted at 3- and 6-months post-enrolment, regardless of whether participants have attended their scheduled clinic visits, to ensure complete longitudinal data capture.

#### Sample size justification

For this longitudinal mixed-methods study, the quantitative sample size was calculated assuming a two-sided significance level (α) of 0.05 and a statistical power of 0.90. Based on an anticipated population standard deviation of 0.55 and an expected difference between the population mean (m) and a specified reference value (m_0_), the minimum required sample size was estimated at 82 participants. This sample size is considered adequate to detect meaningful differences in MHCS engagement over time.

#### Primary quantitative outcome

The primary outcome is mental health care service (MHCS) engagement, measured using the Service Engagement Scale (SES) (22). The SES comprises 14 items assessing four domains: availability, collaboration, help-seeking, and treatment adherence. Total scores range from 0 to 42, with higher scores indicating greater engagement with mental health services.

#### Secondary quantitative outcomes

Secondary outcomes include disability-related indicators (functional impairment, quality of life, and medication side effects), mortality (all-cause mortality and suicide), hospital re-admission, and treatment adherence.

#### Quantitative predictor variables

Predictor variables include duration of untreated psychosis (DUP), use of alternative and complementary therapies, diagnostic category, stigma, symptom profile and severity, cognitive function, physical and metabolic health indicators, medication side effects, and socio-demographic characteristics such as age and sex.

### Qualitative component

The qualitative component will employ a longitudinal qualitative design to explore patient and caregiver lived experiences during the first episode of psychosis and how these experiences shape engagement with mental health care services over time. A participatory visual method, draw–write–and–tell (DWT) will be combined with in-depth interviews to generate rich, participant-led accounts (23).

#### Draw–write–and–tell (DWT) technique

DWT is a participatory visual method that complements traditional qualitative approaches by enabling participants to express experiences, emotions, and meanings that may be difficult to articulate verbally (23, 24). The DWT is particularly suited to FEP contexts, as the technique facilitates reflection on personal and socially situated experiences and supports the exploration of sensitive topics such as illness perceptions, care-seeking trajectories, and treatment experiences. The method enables participants to externalise experiences through visual representation and short written reflections, which are then explored through narrative interviews. Combining visual outputs (drawings and written narratives) with narrative interviews allows participants to guide the discussion, facilitates participant-led meaning-making and supports in-depth exploration of subjective experiences. A longitudinal qualitative design will allow examination of how illness perceptions, care-seeking experiences, and service engagement evolve over time, particularly during the critical early treatment period.

#### Participant selection

Participants for the qualitative component will be purposively selected from individuals enrolled in the quantitative cohort. A maximum variation sampling strategy will be employed to capture a diverse range of experiences across gender, age group, diagnosis, duration of untreated psychosis, and level of service engagement. Approximately 20 patients with FEP will be recruited at baseline. This sample size is informed by the concept of information power, whereby a smaller sample is appropriate when the study aim is focused, the sample is specific, and in-depth longitudinal data are generated. The longitudinal design, involving repeated interviews, is expected to yield substantial analytic depth at the individual case level. In addition, a purposive subsample of caregivers (approximately 15–20) will be recruited to participate in longitudinal in-depth interviews to explore caregiving experiences and perspectives on patient engagement with care. Sampling will remain iterative. Recruitment will continue until sufficient variation is achieved to address the research objectives. This strategy will allow the study to capture a broad range of experiences related to illness onset, care-seeking pathways, and engagement with mental health services, thereby enhancing the depth, richness, and analytical value of the qualitative data.

### Data collection procedures

#### Baseline data collection

At baseline, participants will be provided with a blank A3 sheet of paper and drawing materials (crayons, pencils, or pens) and invited to draw and/or write about their experiences during the onset and early phase of psychosis, including how they understood what was happening, how they sought help, and what engaging with services has meant to them. Participants will be informed that artistic ability is not required and that they may express themselves in any format they prefer. The emphasis will be placed on meaning with no expectation of artistic ability. Following the drawing or writing exercise, participants will take part in an unstructured, in-depth interview. The interview will begin with an open-ended invitation such as: “Can you tell me about what you have drawn/written and what it means to you?” The participant’s narrative will guide the discussion. Interviews will explore; What living with FEP means to them; their recollections of illness onset and early experiences; pathways to care and how they arrived at the mental health care facility; experiences of receiving care at the facility; key challenges or struggles encountered and their hopes and fears for the future. This participant-led approach ensures that the meanings attached to visual outputs remain grounded in participants’ own interpretations. Caregiver interviews will follow a semi-structured format exploring recognition of symptoms, help-seeking decisions, caregiving burden, perceptions of services, and factors influencing sustained engagement. Interviews will be conducted in the participant’s preferred language, audio-recorded with consent, and last approximately 60–90 minutes. Field notes will be documented to capture contextual observations and researcher reflections.

#### Longitudinal follow-up

Participants will be provided with rough sketchbooks and encouraged to continue drawing or writing about their experiences at home between study visits. These materials will serve as reflective tools and prompts for subsequent interviews. Patient and caregiver participants will be re-interviewed at three and six months following the baseline interviews. During these interviews, participants will be invited to reflect on any drawings or writings produced since the previous visit and to discuss changes in their experiences, perceptions of care, and engagement with mental health services over time. Follow-up interviews will explore changes over time in illness perceptions, experiences of treatment and service engagement, adherence and discontinuation, social support and stigma, functional recovery and future outlook. This process will support exploration of evolving illness narratives and trajectories of engagement.

This longitudinal approach will enable the study to capture evolving experiences and meanings associated with illness and service engagement during the early course of psychosis.

The qualitative findings will provide contextualised, in-depth insights into how patients and caregivers understand FEP, navigate care pathways, and experience mental health services. When integrated with quantitative findings, this component will strengthen interpretation of service engagement outcomes and inform the development of more responsive and patient-centred mental health interventions.

#### Retention Strategy

We will utilize various strategies to reduce loss to follow-up, a significant problem in this population (2). First, phone follow-ups by research assistants will remind discharged patients of interviews (25). Furthermore, including family members will improve retention as family involvement strongly predicts a return to care (6).

### Data analysis

This study adopts a longitudinal convergent mixed-methods design embedded within a prospective cohort. This integrative approach will strengthen explanatory depth and enhance the service and policy relevance of findings in the Ugandan mental health context.

#### Quantitative data analysis

Baseline characteristics of participants will be summarised using means and standard deviations (or medians and interquartile ranges for skewed distributions) for continuous variables, and frequencies and percentages for categorical variables. Potential confounders will be identified a priori, including age, sex, and welfare status, and will be adjusted for in the main outcome analysis. The primary outcome is mental health care service (MHCS) engagement, measured using the Service Engagement Scale (SES), calculated as the mean score across the three- and six-month follow-up visits. Secondary outcomes include functional impairment, quality of life (assessed using WHOQOL-BREF), medication adherence, re-admission, and all-cause mortality. Covariates will include sociodemographic, clinical, welfare, stigma, economic, and functional variables. To examine changes over time, random-effects linear regression models will be used for continuous outcomes, accounting for repeated measures within participants. Sensitivity analyses will be conducted to assess the impact of missing data. All analyses will be conducted using Stata version 18, and statistical significance will be set at p < 0.05.

#### Qualitative data analysis

Qualitative data will include visual artefacts (drawings and written reflections), interview transcripts, and field notes. Interpretative Phenomenological Analysis (IPA) will be used to explore participants lived experiences during the first episode of psychosis. Audio recordings will be transcribed verbatim and translated into English where necessary. Transcripts will be checked against audio files for accuracy. Transcripts will be analysed alongside drawings and written materials from the Draw–Write–and–Tell (DWT) exercises, considering aspects such as style, tone, colour, and content. Written reflections and narratives embedded in the drawings will be integrated to generate in-depth, idiographic accounts of participants’ experiences. Themes will be generated within these groups to explore how lived experiences relate to engagement patterns, while remaining grounded in participants’ own accounts. Longitudinal comparisons of drawings, writings, and interview data across baseline, three, and six months will be conducted to examine changes in experiences, perceptions, and engagement over time.

#### Rigour and trustworthiness

Analyst triangulation will be employed, with at least two researchers independently reviewing selected transcripts and visual materials. Reflexive team discussions will be conducted to examine interpretive assumptions. An audit trail will be maintained documenting analytic decisions. Visual artefacts will not be interpreted independently of participants’ narratives; meaning attribution will remain grounded in participants’ explanations.

#### Triangulation of quantitative and qualitative data

A convergent mixed-methods approach will be adopted to integrate quantitative and qualitative findings. Quantitative data on MHCS engagement and covariates will be triangulated with qualitative themes to develop a comprehensive understanding of patient and caregiver experiences. This approach will allow identification of patterns and mechanisms underlying engagement. This understanding allows new theoretical propositions about engagement to be developed for future implementation studies and interventions in mental health service delivery.

### Data management plan and quality control

#### Quantitative data

Data will be captured electronically using Open Data Kit (ODK), ensuring real-time uploading and secure storage on a password-protected server. Each participant’s assessments will be linked to a unique identifier to consolidate all data within the same folder. Datasets will be anonymised prior to analysis and securely backed up on Mendeley. Access to electronic data will be restricted to the core research team, all of whom will have received training in study procedures and completed certifications in ethical research conduct. Data integrity will be ensured through regular checks for completeness, consistency, and accuracy.

#### Qualitative data

All interviews and Draw-Write-and-Tell (DWT) sessions will be audio-recorded using encrypted digital recorders. Audio files will be transferred daily to a secure, password-protected server and backed up on a second encrypted drive. Drawings and written materials from DWT exercises will be scanned and stored as PDF or image files in the same secure folder linked to the participant’s unique identifier.

Audio recordings will be transcribed verbatim, with transcripts checked for accuracy by a second team member. Drawings/writings will be coded and catalogued systematically, ensuring they are linked to the correct participant while maintaining anonymity. Identifiable information will be removed from transcripts and images prior to analysis.

#### Access and Oversight

Only core research team members will have access to the qualitative and quantitative data. All team members will have training in ethical research conduct, including confidentiality and secure data handling. Any paper-based notes collected during interviews will be stored in locked cabinets, accessible only to authorised personnel.

#### Quality Assurance

To maintain rigour, all qualitative coding will be conducted in duplicate, with discrepancies resolved through team discussion. Regular audit checks will verify that transcripts, drawings, and recordings are complete, anonymised, and stored according to protocol. This ensures integrity, confidentiality, and reproducibility of both quantitative and qualitative data.

#### Study governance, stakeholder engagement, and community oversight

A governance council will oversee all study activities to ensure ethical, safe, and contextually appropriate conduct alongside the research team. The council will be led by lived-experience experts from Heart to Heart Spaces, https://hearttoheartspaces.com/; a mental health community providing safe spaces for individuals to share their experiences, coping mechanisms, and recovery strategies. The council will review the study protocol and data collection tools prior to study initiation and observe the piloting of procedures to suggest refinements aimed at improving data quality, participant engagement, and retention. Heart to Heart Spaces has been involved in the design of the protocol and will continue to advise on participant engagement and the ethical conduct of the study. The council ensures the study aligns with community perspectives, promoting accountability, transparency, and ethical responsiveness throughout the research process.

#### Ethics

Ethical approval has been obtained from the School of Medicine Research and Ethics Committee (SOMREC) of Makerere University Ref: Mak-SOMREC-2024-1002 and administrative approval from the Butabika Hospital Institutional Review Board. Informed consent will be obtained before any study procedures. The potential participant will ask questions before providing a signed or thumbprint consent upon demonstrating an understanding of the study procedures. Study participants will receive a financial incentive to defray transport costs.

#### Assessing the capacity to consent and informed consent

The University of California, San Diego Brief Assessment of Capacity to Consent (UBACC) instrument will assess whether the participants have understood the consent process, including the risks and benefits of the study (15). It is a 10-item scale comprised of 3 factors that evaluate understanding, appreciation and reasoning. Scores of less than 14.5 on three occasions indicate that the participant has not understood and will not be enrolled. The UBACC is translated to Luganda and previously used in this setting (15). Given the vulnerability of individuals experiencing FEP, capacity to provide informed consent will be assessed in collaboration with the clinical team prior to participation. Participation will occur only when individuals are clinically stable and able to provide informed consent. Participants may pause or withdraw at any time. If distress arises during the DWT exercise or interviews, the session will be paused and, if necessary, participants will be referred to the attending clinical team for support. Interviews will be conducted in private spaces within the facility to ensure confidentiality.

#### Confidentiality and access to data

All study data, including quantitative assessments, qualitative interview recordings, and participant drawings/writings, will be securely stored on password-protected servers and linked to unique participant IDs. Only the core research team will have access, and all identifiers will be removed from datasets used for analysis. No individual-level data will be included in publications or presentations.

#### Dissemination policy

Study findings will be disseminated through peer-reviewed journal publications, conference presentations, and community reports. Patients, caregivers, and the Heart to Heart Spaces community will be involved in sharing results in formats accessible to service users. The study team will retain full independence in deciding the content and timing of publications, without involvement of any external sponsor.

#### Data availability

Deidentified quantitative and qualitative data will be made available to researchers upon reasonable request, in line with ethical approvals and participant consent. Qualitative drawings and writings will be shared in a de-identified form to protect participant confidentiality.

#### Patient and public involvement

The study was developed in collaboration with *Heart to Heart Spaces*, a mental health community of lived-experience experts, who contributed to the study design and will serve on the governance committee to protect patient interests. They will also support participant follow-up and dissemination of study findings to ensure that outcomes are relevant and accessible to service users. A subset of patient and caregiver participants will be invited to provide feedback on preliminary qualitative findings to enhance interpretive validity and ensure that themes accurately reflect lived experiences. At the dissemination stage, findings will be shared with patient and caregiver groups through stakeholder meetings at the study site. A plain-language summary of findings will be developed and made available to participants and community stakeholders. The research team will also engage with *Heart to Heart Spaces* to support broader dissemination and translation of findings into practice.

## Discussion

This longitudinal mixed-methods study will explore patient and caregiver lived experiences and their relationship to engagement with mental healthcare services (MHCS) during first-episode psychosis in Uganda. By combining quantitative assessment of service engagement with participatory qualitative methods, including the draw-write-and-tell technique, the study will provide nuanced insights into factors influencing engagement, such as stigma, alternative treatment-seeking, and systemic barriers.

Engaging both patients and caregivers, and incorporating lived-expert perspectives through Heart to Heart Spaces, strengthens the study’s relevance and aims to reduce attrition, a common challenge in first-episode psychosis research. Triangulation of qualitative and quantitative data will support a comprehensive understanding of patterns of disengagement and re-engagement, enabling identification of actionable strategies to enhance early intervention, continuity of care, and patient-centered decision-making.

While the study is limited by its single-site design and the exclusion of patients with comorbid substance use disorders, these measures help focus on MHCS engagement in a defined population and reduce clinical heterogeneity. Findings from this study will inform culturally relevant interventions to improve service access, treatment adherence, and early support for patients and caregivers. Other potential limitations include the single-site design, which may limit generalizability beyond the Greater Kampala area, and the exclusion of patients with comorbid substance use disorders, which may affect the representativeness of findings. Nevertheless, embedding lived-expert input and systematic follow-up strategies aims to minimize attrition and enhance the validity and relevance of the study findings

## Conclusion

This protocol outlines a novel approach to understanding the lived experiences of patients and caregivers during first-episode psychosis and their engagement with mental health services. By combining longitudinal quantitative assessment with participatory qualitative methods, the study will generate evidence to inform the design of interventions that enhance early engagement, improve treatment adherence, and support recovery. The findings will provide actionable insights for clinicians, policymakers, and community stakeholders to optimize mental health service delivery in Uganda and similar low-resource settings.

## Data Availability

All data produced in the present work will be contained in the manuscript

## Author Contribution

PO and ASS drafted the manuscript and are joint first authors. EKM, ASS, WS, EN and WM contributed to the conceptualisation of the study design. PO, ASS, WS, RA, SB, BTA, AKK, MA, EKM, are members of the study team who have contributed to specifying the study design. All coauthors have revised the manuscript critically for important intellectual content.

## References

1. Anderson KK, Norman R, MacDougall A, Edwards J, Palaniyappan L, Lau C, et al. Effectiveness of Early Psychosis Intervention: Comparison of Service Users and Nonusers in Population-Based Health Administrative Data. Am J Psychiatry. 2018;175(5):443–52.

2. Mwesiga E, Nakasujja N, Ongeri L, Loewy R, Meffert S, Semeere A. Trends and Predictors of Engagement with Mental Healthcare Services in the Year Following Initiation of Antipsychotic Medication Among Ugandan First-Episode Psychosis Patients. SSRN Electronic Journal. 2022.

3. Lal S, Malla A. Service Engagement in First-Episode Psychosis: Current Issues and Future Directions. Can J Psychiatry. 2015;60(8):341–5.

4. Casey D, Brown L, Gajwani R, Islam Z, Jasani R, Parsons H, et al. Predictors of engagement in first-episode psychosis. Schizophr Res. 2016;175(1-3):204–8.

5. Myers N, Bhatty S, Broussard B, Compton MT. Clinical Correlates of Initial Treatment Disengagement in First-Episode Psychosis. Clin Schizophr Relat Psychoses. 2017;11(2):95–102.

6. Mwesiga EK, Ssemata AS, Nakitende AJ, Ongeri L, Semeere A, Loewy R, et al. Exploring facilitators for a transition from alternative and complementary therapies to evidence-based treatments in Ugandan first-episode psychosis patients. medRxiv. 2022:2022.02.21.22270378.

7. Akena D, Semeere A, Kadama P, Mwesiga E, Basangwa D, Nakku J, et al. Clinical outcomes among individuals with a first episode psychosis attending Butabika National Mental Referral Hospital in Uganda: a longitudinal cohort study. A study protocol for a longitudinal cohort study. BMJ Open. 2020;10(6):e034367.

8. Mascayano F, van der Ven E, Martinez-Ales G, Henao AR, Zambrano J, Jones N, et al. Disengagement From Early Intervention Services for Psychosis: A Systematic Review. Psychiatr Serv. 2021;72(1):49–60.

9. Yang LH, Blasco D, Lieff SA, L. PD, Li Y, Broeker M, et al. Stigma of Treatment Stages for First-Episode Psychosis: A Conceptual Framework for Early Intervention Services. Harv Rev Psychiatry. 2021;29(2):131–41.

10. Connor C, Greenfield S, Lester H, Channa S, Palmer C, Barker C, et al. Seeking help for first-episode psychosis: a family narrative. Early Interv Psychiatry. 2016;10(4):334–45.

11. Creswell JW, Klassen AC, Plano Clark VL, Smith KC. Best practices for mixed methods research in the health sciences. Bethesda (Maryland): National Institutes of Health. 2011;2013(2011):541–5.

12. Tindall R, Simmons M, Allott K, Hamilton B. Disengagement Processes Within an Early Intervention Service for First-Episode Psychosis: A Longitudinal, Qualitative, Multi-Perspective Study. Front Psychiatry. 2020;11:565.

13. Lucksted A, Essock SM, Stevenson J, Mendon SJ, Nossel IR, Goldman HH, et al. Client Views of Engagement in the RAISE Connection Program for Early Psychosis Recovery. Psychiatr Serv. 2015;66(7):699–704.

14. Mwesiga EK. Neuropsychological assessment for first-episode psychosis patients in low resource settings 2021.

15. Mwesiga EK, Nakasujja N, Nakku J, Nanyonga A, Gumikiriza JL, Bangirana P, et al. One year prevalence of psychotic disorders among first treatment contact patients at the National Psychiatric Referral and Teaching Hospital in Uganda. PloS one. 2020;15(1):e0218843.

16. Akena D, Semeere A, Kadama P, Mwesiga EK. Feasibility of conducting a pilot randomized control trial of a psycho-education intervention in patients with a first episode psychosis in Uganda-A study protocol. 2022;17(7):e0268493.

17. Mwesiga EK, Nakasujja N, Nankaba L, Nakku J, Musisi S. Quality of individual and group level interventions for first-episode psychosis at the tertiary psychiatric hospital in Uganda. S Afr J Psychiatr. 2021;27:1604.

18. Lecrubier Y, Sheehan D, Hergueta T, Weiller E. The mini international neuropsychiatric interview. European Psychiatry. 1998;13(1004):198s–s.

19. Sheehan D, Janavs J, Sheehan K, Sheehan M, Gray C. Mini international neuropsychiatric interview 6.0: High prevalence disorders. English version Tampa, FL[Google Scholar]. 2010.

20. Breitborde NJ, Srihari VH, Woods SW. Review of the operational definition for first-episode psychosis. Early intervention in psychiatry. 2009;3(4):259–65.

21. GarcÍa S, Martinez-Cengotitabengoa M, Lopez-Zurbano S, Zorrilla I, Lopez P, Vieta E, et al. Adherence to antipsychotic medication in bipolar disorder and schizophrenic patients: a systematic review. Journal of clinical psychopharmacology. 2016;36(4):355–71.

22. Tait L, Birchwood M, Trower P. A new scale (SES) to measure engagement with community mental health services. J Ment Health. 2002;11(2):191–8.

23. Angell C, Alexander J, Hunt JA. ‘Draw, write and tell’: A literature review and methodological development on the ‘draw and write’research method. Journal of early childhood research. 2015;13(1):17–28.

24. Maynes MJ, Pierce JL, Laslett B. Telling stories: The use of personal narratives in the social sciences and history: Cornell University Press; 2012.

25. D’Arcey J, Collaton J, Kozloff N, Voineskos AN, Kidd SA, Foussias G. The use of text messaging to improve clinical engagement for individuals with psychosis: systematic review. JMIR mental health. 2020;7(4):e16993.

